# Suboptimal SARS-CoV-2-specific CD8^+^ T-cell response associated with the prominent HLA-A*02:01 phenotype

**DOI:** 10.1101/2020.08.17.20176370

**Authors:** Jennifer R Habel, Thi H O Nguyen, Carolien E van de Sandt, Jennifer A Juno, Priyanka Chaurasia, Kathleen Wragg, Marios Koutsakos, Luca Hensen, Xiaoxiao Jia, Brendon Chua, Wuji Zhang, Hyon-Xhi Tan, Katie L Flanagan, Denise L Doolan, Joseph Torresi, Weisan Chen, Linda M Wakim, Allen C Cheng, Peter C Doherty, Jan Petersen, Jamie Rossjohn, Adam K Wheatley, Stephen J Kent, Louise C Rowntree, Katherine Kedzierska

## Abstract

An improved understanding of human T-cell-mediated immunity in COVID-19 is important if we are to optimize therapeutic and vaccine strategies. Experience with influenza shows that infection primes CD8^+^ T-cell memory to shared peptides presented by common HLA types like HLA-A2. Following re-infection, cross-reactive CD8^+^ T-cells enhance recovery and diminish clinical severity. Stimulating peripheral blood mononuclear cells from COVID-19 convalescent patients with overlapping peptides from SARS-CoV-2 Spike, Nucleocapsid and Membrane proteins led to the clonal expansion of SARS-CoV-2-specific CD8^+^ and CD4^+^ T-cells *in vitro*, with CD4^+^ sets being typically robust. For CD8^+^ T-cells taken directly *ex vivo*, we identified two HLA-A*02:01-restricted SARS-CoV-2 epitopes, A2/S_269–277_ and A2/Orf1ab_3183–3191_. Using peptide-HLA tetramer enrichment, direct *ex vivo* assessment of the A2/S_269_^+^CD8^+^ and A2/Orf1ab_3183_^+^CD8^+^ populations indicated that the more prominent A2/S_269_^+^CD8^+^ set was detected at comparable frequency (∼1.3×10^−5^) in acute and convalescent HLA-A*02:01^+^ patients. But, while the numbers were higher than those found in uninfected HLA-A*02:01^+^ donors *(∼*2.5×10^−6^), they were low when compared with frequencies for influenza-specific (A2/M1_58_) and EBV-specific (A2/BMLF_1280_) *(∼*1.38×10^−4^) populations. Phenotypic analysis *ex vivo* of A2/S_269_^+^CD8^+^ T-cells from COVID-19 convalescents showed that A2/S_269_^+^CD8^+^ T-cells were predominantly negative for the CD38, HLA-DR, PD-1 and CD71 activation markers, although the majority of total CD8^+^ T-cells were granzyme and/or perforin-positive. Furthermore, the bias towards naïve, stem cell memory and central memory A2/S_269_^+^CD8^+^ T-cells rather than effector memory populations suggests that SARS-CoV2 infection may be compromising CD8^+^ T-cell activation. Priming with an appropriate vaccine may thus have great value for optimizing protective CD8^+^ T-cell immunity in COVID-19.

## INTRODUCTION

The current SARS-CoV-2 pandemic has, as of August 2020, infected more than 21 million people, caused at least 700,000 deaths (1) and paralysed economies globally. Although the majority of infections are mild-to-moderate and short in duration, ∼12–18% of COVID-19 patients develop severe disease requiring hospitalization, ∼5% are critical (2–4), and others who are less severely affected, and even asymptomatic, may still have some underlying pathology (5). These are still early days, and there is much that remains unknown about both the innate and adaptive immune responses in COVID-19. An urgent need is to develop a better understanding so that any immunopathology can be managed, and vaccine design and immunotherapies optimized.

So far as adaptive immunity is concerned, we do know that SARS-CoV-2-specific antibodies can be found in ∼95% of convalescent COVID-19 patients (6, 7) and that titres determined in virus neutralization assays correlate well with spike-protein-binding immunoglobulin (Ig) levels measured by ELISA (8, 9). High serum neutralizing antibody titers tend to be more prominent in severe COVID-19, which could be characteristic of prolonged antigen stimulation due to delayed virus clearance. Otherwise, the duration of SARS-CoV2-specific IgG persistence in serum is far from clear, and we have much to learn about the CD4^+^ and CD8^+^ T-cell responses.

Virus-specific CD8^+^ T-cells are generally thought to be involved in the elimination of virus-infected cell ‘factories’ in the acute response to respiratory viruses with, where there is established CD8^+^ T-cell memory, that response being enhanced in both rapidity and magnitude to provide a measure of protection against the development of severe disease following secondary virus challenge. Survivors of the 2002–3 SARS outbreak still maintain CD4^+^ and CD8^+^ T-cell populations reactive to the SARS-CoV-1 nucleocapsid protein (10) and evidence of sustained T-cell memory has also been found for MERS (11). Furthermore, it is possible that there may be some cross-reactive T-cell memory for COVID-19 in people who have been infected with these viruses and, perhaps, more broadly, with the previously circulating common cold coronaviruses (12).

For SARS-CoV-2 there is growing evidence that virus-specific T-cells are indeed being generated. Our early COVID-19 case study showed that both CD4^+^ T-follicular helper cells and activated CD38^+^HLA-DR^+^CD8^+^ T-cells appeared in patient’s blood at 3 days prior to recovery, suggesting that they played a part in the resolution of COVID-19 (13). Recent communications from others also reported the presence SARS-CoV-2-reactive CD4^+^ and CD8^+^ T-cells in both acute and convalescent COVID-19 patients (14, 15). More disturbing is, however, an analysis suggesting that at least a proportion of the SARS-CoV-2-specific CD8^+^ T-cells recovered from peripheral blood may be showing as an ‘exhausted’ phenotype (16). Clearly, it is a matter of urgency to develop a better understanding of the integrity of the acute CD8^+^ T-cell response in COVID-19 and how this impacts on disease outcome.

Here, we utilized a combination of peptide prediction and *in vitro* peptide stimulation with overlapping peptides from the Spike, Nucleocapsid and Membrane proteins to identify two novel SARS-CoV2 epitopes restricted by HLA-A*02:01 (A2/S_269_ and A2/Orf1ab_3183_) in individuals with COVID-19. Using peptide-HLA-I tetramers, we performed direct *ex vivo* tetramer enrichment to define the frequency and activation profiles of the responding SARS-CoV2-specific CD8^+^ T-cells in acute and convalescent COVID-19 patients and in pre-pandemic PBMCs, tonsil and lung tissues from uninfected donors.

Our data establish that HLA-A*02:01-restricted SARS-CoV-2-reactive CD8^+^ T-cells can be detected directly *ex vivo* in both COVID-19 patients and in immunologically naïve individuals. However, while SARS-CoV-2-specific CD4^+^ T-cell responses were broadly comparable to those found previously for other viruses, virus-activated CD8^+^ T-cells that recognize SARS-CoV2 peptides presented by the common (at least in Caucasians) HLA-A*02:01 MHC-I glycoprotein were both at low prevalence and express a less than optimal (for virus elimination) phenotype. These findings raise a number of questions. Is this apparent CD8^+^ T-cell response defect limited to these particular epitopes? If so, are HLA-A*02:01 individuals at higher relative risk? Alternatively, if this is a general effect, is the SARS-CoV-2 virus in some way subverting CD8^+^ T-cell responsiveness? Perhaps COVID-19 may be one disease where an appropriately designed vaccine may do better than nature when it comes to generating a protective CD8^+^ T cell recall response.

## RESULTS

### COVID-19 patient cohort and uninfected controls

This study of 18 COVID-19 cases included one person who remained asymptomatic, 10 who were symptomatic but were cared for at home, and 7 who were admitted to hospital including 2 requiring supplemental oxygen (Supplementary Table 1). Control cells were tested from another 17 uninfected individuals who formed a control group (Supplementary Table 2). All COVID-19 patients (median age 54 years, 55.6% females) seroconverted for SARS-CoV-2 antibodies by Receptor Binding Domain (RBD) ELISA (17) and 12 were HLA-A*02:01-expressing individuals. As controls, we analysed pre-existing A2/CD8^+^ T-cell responses in pre-pandemic PBMC and tonsil samples from 12 HLA-A*02:01-expressing subjects across three age groups: children (median age 9.5 years), adults (median age 51 years) and the elderly (median age 66.5 years) with 44% being female (Supplementary Table 2). Additionally, we tested pre-existing A2/CD8^+^ T-cell populations in lung tissues from 5 HLA-A2 individuals (median age 42 years).

### CD4^+^ and CD8^+^ T-cell responses to SARS-CoV-2 overlapping peptide pools

We first probed for SARS-CoV-2-specific CD4^+^ and CD8^+^ T-cells in convalescent COVID-19 donors using a standard 6-hr intracellular cytokine staining (ICS) assay using peptide pools containing 15mers, overlapping by 11 amino acids, which spanned the entire Nucleocapsid(N) and Membrane (M) proteins and selected regions of SARS-CoV2 Spike (S) protein. The PBMCs were stimulated with one peptide pool and expanded for 10 days before the assessment of SARS-CoV-2-reactive T-cells by ICS for intracellular IFN-_γ_, TNF and MIP-1β, plus staining for CD107a and perforin (Fig. 1; Supplementary Fig 1A) using individual peptide pools. The responding CD4^+^ T-cells all stained for IFN-_γ_, TNF, MIP-1β, CD107a and perforin, while the CD8^+^ T-cells were predominately positive for perforin (Fig 1AB). The CD4^+^ T-cells showed significant staining for IFN-_γ_, with 5/6 subjects generating IFN-_γ_^+^CD4^+^ T-cells responses to at least one of the SARS-CoV-2 peptide N, M or S pools, indicating that convalescent COVID-19 patients have solid SARS-CoV-2 specific CD4^+^ T-cell immunity. However, while CD8^+^ T cells from 3/6 donors were perforin-positive, evidence of modest IFN-_γ_^+^ activation for the CD8^+^ set was found in only 1/6 donor. It thus seems that IFN-_γ_-producing SARS-CoV2 specific CD4^+^ T-cells expand to a much greater extent than the CD8^+^ set following *in vitro* peptide stimulation (Fig. 1C).

**Fig 1.**
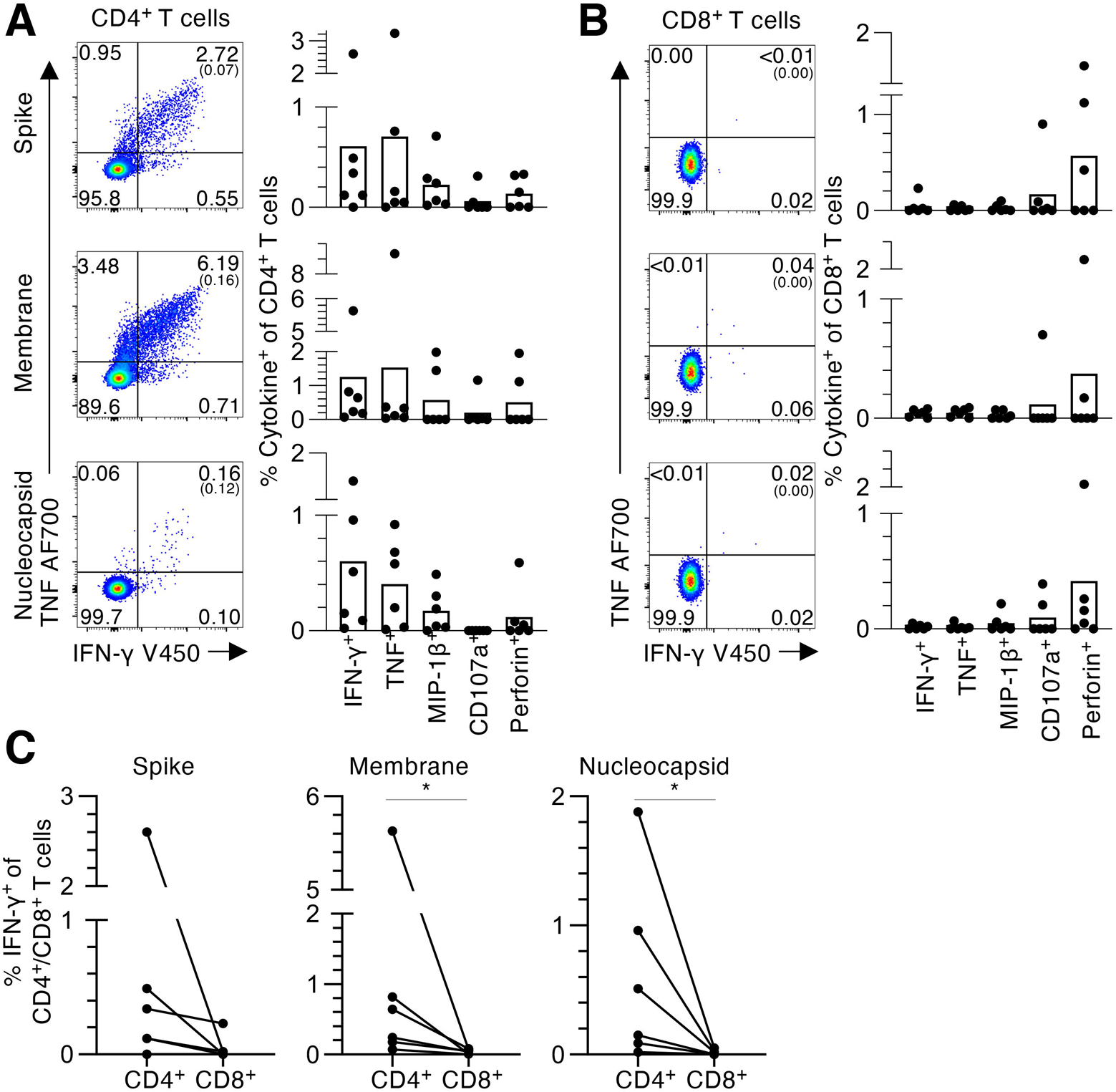
CD4^+^ and CD8^+^ T-cell responses to SARS-CoV-2 overlapping peptide pools. (A) CD4^+^ and (B) CD8^+^ T-cell responses to SARS-CoV2 spike (S), membrane (M) and nucleocapsid (N) peptide pools in convalescent COVID-19 individuals. (i) Representative FACS plots showing IFN-_γ_ and TNF staining of (A) CD4^+^ or (B) CD8^+^ T cell populations. (ii) Frequencies of IFN-_γ_^+^, TNF^+^, MIP-1β^+^, CD107a^+^ or perforin^+^ within the (A) CD4^+^ or (B) CD8^+^ T-cells, with background staining subtracted (n = 6, mean). Background staining values are shown in brackets. Data points show individual COVID-19 convalescent subjects. (C) Paired frequencies of IFN-_γ_^+^CD4^+^ and CD8^+^ T-cells for S, N, and M peptide pools. Statistical significance was determined with Wilcoxon matched-pairs signed rank test, \**P*< 0.05.

### Identification of SARS-CoV-2-specific HLA-A*02:01-restricted CD8^+^ T-cell epitopes

Switching the focus to HLA-specific SARS-CoV-2 CD8^+^ T-cell responses, we next identified CD8^+^ T-cell specificities for HLA-A*02:01-expressing individuals. Using predicted HLA-A*02:01-binding SARS-CoV2-derived peptides from the SARS-CoV-2 S, N, M and Polyprotein1ab (Orf1ab) proteins (Supplementary Table 3; based on two prediction algorithms: NetCTLpan and NetMHCpan; accessed 27 March 2020), PBMCs from 5 convalescent HLA-A*02:01^+^ COVID-19 individuals were expanded *in vitro* with a pool of 14 predicted A2/SARS-CoV2 peptides for 10 days, then restimulated with individual peptides in an ICS assay to determine peptide immunogenicity. Of the 14 peptides screened, S_269–277_ (YLQPRTFLL) generated the strongest CD8^+^IFN-_γ_^+^response (mean 0.19%, n = 5), with lesser responses being elicited for S_976–984_ (VLNDILSRL) and Orf1ab_3183–3191_ (FLLNKEMYL) (0.07% and 0.08%, respectively, mean, n = 5) (Fig 2; Supplementary Fig 1B). Collectively, we identified one dominant and two subdominant, novel A2/CD8^+^ T cell specificities for COVID-19.

**Fig 2.**
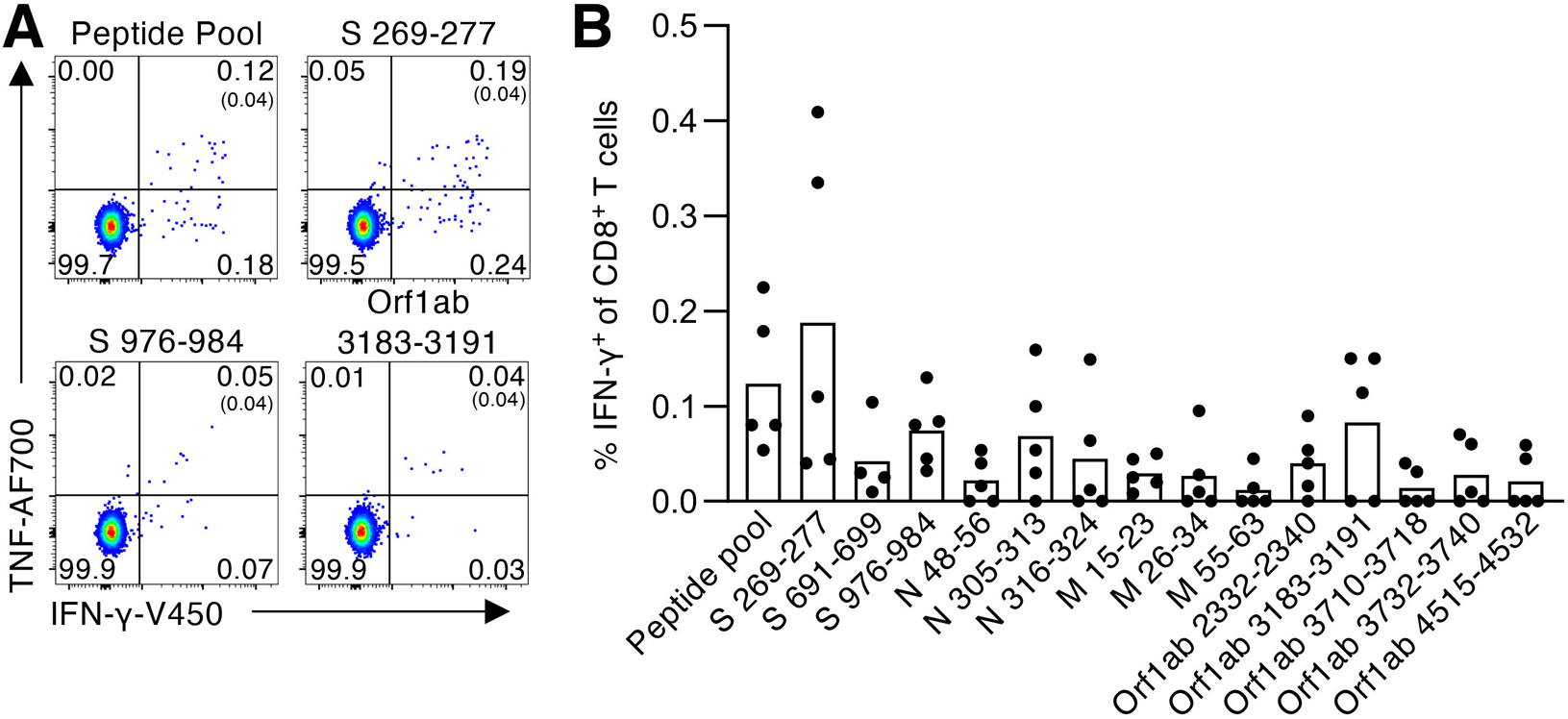
Identification of SARS-CoV-2-specific HLA-A*02:01-restricted CD8^+^ T-cell epitopes. (A) Representative FACS plots of CD8^+^ IFN-_γ_/TNF staining after stimulation with the SARS-CoV-2 predicted peptide pool and individual S_269–277_, S_976–984_ and Orf1ab_3183–3191_ peptides. (B) Frequency of IFN-_γ_^+^ of CD8^+^ T-cells to each SARS-CoV-2 peptide within the predicted peptide pool, with background staining subtracted (n = 5, mean). Peptide screen was performed in convalescent COVID-19 PBMCs after 10-day expansion *in vitro*.

Peptide sequence conservation analysis for these SARS-CoV-2 immunogenic peptides was extended to previously circulating coronaviruses. Reference protein sequences for SARS-CoV-1 and MERS plus the ‘common cold’ human CoV (hCoV) strains 229E, HKU1, NL63 and OC43 were obtained from NCBI. Using the Virus Pathogen Resource (http://www.viprbrc.org), SARS-CoV2 S_269,_ S_976_ and Orf1ab_3183_ peptide sequences were compared to their respective protein sequences within each CoV strain (Supplementary Table 3). Our data showed that SARS-CoV2/Orf1ab_3183_ and S_976_ lacked any sequence similarity to hCoV or MERS strains, but each shared 100% sequence identity with SARS-CoV1/Orf1ab_3160–3168_ (FLLNKEMYL) and S_958–966_ (VLNDILSRL), respectively. SARS-CoV-2/S_269_ shared 78% and 67% sequence identity with MERS/S_317–325_ (<u>K</u>LQP<u>L</u>TFLL) and SARS-CoV-1/S_256–264_ (YL<u>K</u>P<u>T</u>TF<u>M</u>L), respectively. Evidently the A2/SARS-CoV-2 CD8^+^ T-cell epitopes identified may be cross-reactive for SARS-CoV-1 and MERS, while they did not share homology with the common cold hCoVs that circulate in Australia.

### SARS-CoV-2-specific A2/CD8^+^ T-cells are at low frequency in COVID-19 patients

To further analyse the SARS-CoV-2-specific A2/CD8^+^ populations from COVID-19 patients, we generated tetramers for the A2/S_269_ and A2/Orf1ab_3183_ epitopes. Tetramer-associated magnetic enrichment (18, 19) was then used to determine the *ex vivo* frequencies of A2/S_269_^+^CD8^+^ and A2/Orf1ab_3183_^+^CD8^+^ T-cells in acute and convalescent HLA-A*02:01^+^ cases. During the acute phase of COVID-19, A2/S_269_^+^CD8^+^ T-cells were readily detected after *ex vivo* tetramer enrichment at a mean frequency of 1.44×10^−5^ (n = 3) in the CD8^+^ set, while the values for the A2/S_269_^+^CD8^+^ and A2/Orf1ab_3138_^+^CD8^+^ T-cells from COVID-19 convalescents were 1.28×10^−5^ (n = 14) and 1.77×10^−6^ (n = 6), respectively (Fig 3AD). There was no significant difference in the frequency of A2/S_269_^+^CD8^+^ T-cells between acute and convalescent COVID-19 donors, while minimal A2/S_269_^+^CD8^+^ and A2/Orf1ab_3183_^+^CD8^+^ T-cells were detected in either unenriched or flow-through samples (Supplementary Fig 2). Indeed, while too few T-cells were available to test other specificities concurrently for the COVID-19 patients, these frequencies of SARS-CoV-2-specific CD8^+^ T-cells were significantly lower than those found for influenza A virus (IAV)-specific (1.39×10^−4^ for A2/M1_58_; n = 6) and EBV-specific (1.38 ×10^−4^ for A2/BMLF_1280_; n = 6) memory T cell populations from uninfected controls (Fig 3BD), and as per previous publications (19, 20).

**Fig 3.**
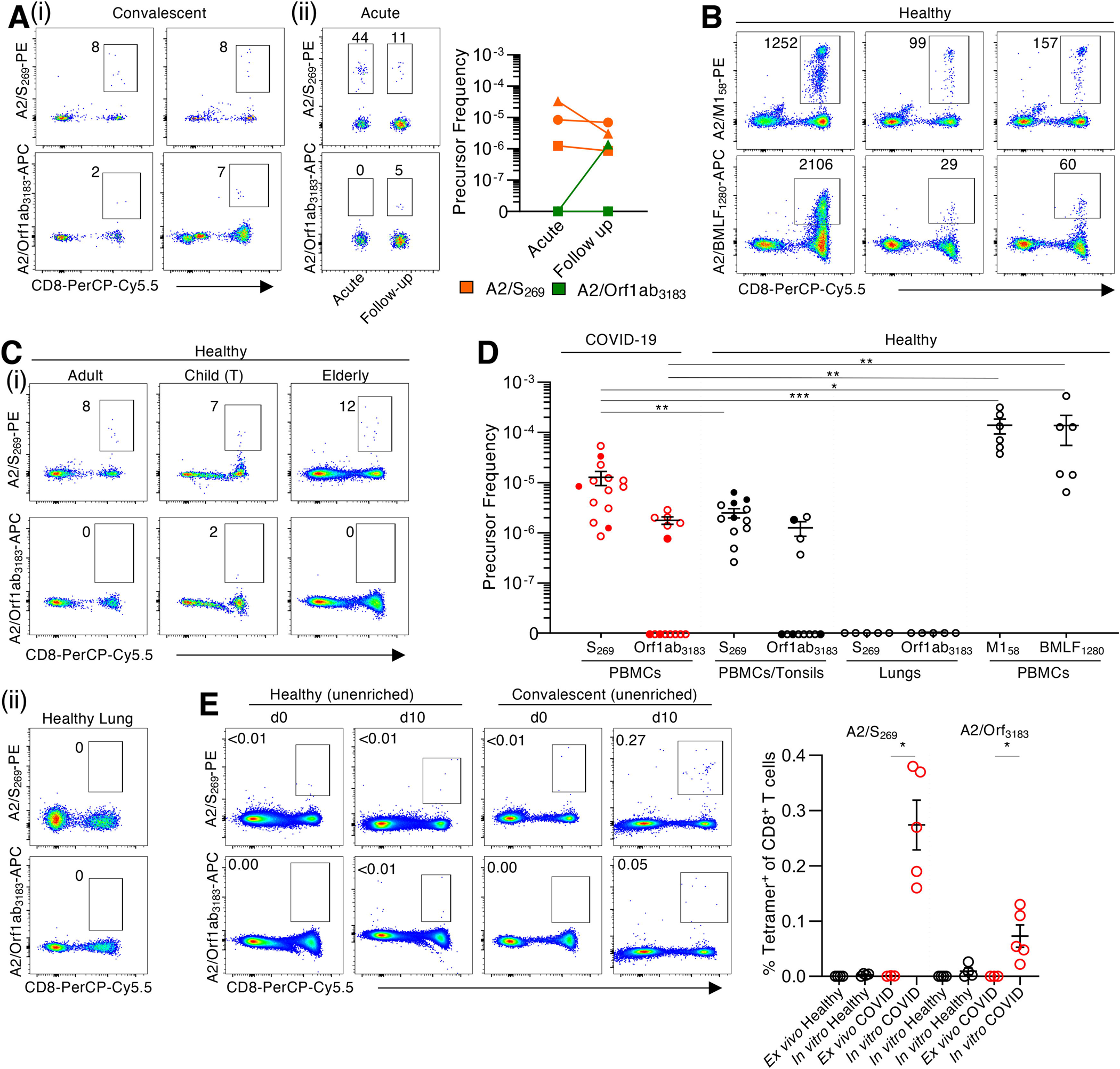
Low *ex vivo* frequency of SARS-CoV-2-specific A2/CD8^+^ T-cell specificities in acute and convalescent COVID-19 patients. A2/S_269_^+^CD8^+^ and A2/Orf1ab_3183_^+^CD8^+^ T cells were identified directly *ex vivo* from healthy (pre-COVID-19) PBMCs, tonsils and lungs; as well as acute and convalescent COVID-19 PBMCs by tetramer magnetic enrichment. (A) Representative FACS plots of A2/S_269_^+^CD8^+^ and A2/Orf1ab_3183_^+^CD8^+^ T-269 3183 cells from enriched samples of (i) convalescent and (ii) acute COVID-19 PBMCs. (B) Representative FACS plots of A2/M1_58_^+^CD8^+^ and A2/BMLF_1280_^+^CD8^+^ T-cells from enriched healthy PBMCs. (C) Representative FACS plots of A2/S_269_^+^CD8^+^ and A2/Orf1ab_3183_^+^CD8^+^ T-cells from enriched adult and elderly PBMCs, and children tonsils (T). (D) A2/CD8^+^ T-cells precursor frequencies were calculated for A2/S_269_^+^CD8^+^, A2/Orf1ab_3183_^+^CD8^+^,A2/M1_58_^+^CD8^+^ and A2/BMLF_1280_^+^CD8^+^ T-cells enriched from either PBMCs or tonsils, or stained in lungs. Dots represent individual donors. Means±SEM are shown. Red dots are COVID-19 acute (closed circle) and convalescent (open circle) donors. Black dots are healthy adult or elderly PBMCs (open circle), or MNCs from child tonsils (closed circle). Donors with undetectable precursor frequencies of 0 are included on the graph to show the number of donors tested. These donors were not included in statistical analyses. Statistical significance was determined with two-tailed Mann-Whitney test, \**P*<0.05, \*\**P*<0.01, \*\*\**P*< 0.001. (E) Representative FACS plots and frequencies of A2/SARS-CoV-2^+^CD8^+^ T cells in the CD8^+^ population in healthy and convalescent donors on d0 and d10 of expansion. Dots represent individual donors. Statistical significance was determined using a two-tailed Mann Whitney test, \**P*< 0.05, \*\**P*< 0.01, \*\*\**P*< 0.001.

Are SARS-CoV-2-specific CD8^+^ T-cells present in uninfected people? Using *ex vivo* tetramer enrichment with pre-pandemic PBMC, tonsil and lung samples taken from HLA-A*02:01-expressing unifected individuals (Fig. 3CiD), naïve SARS-CoV-2-specific CD8^+^ T-cells directed at A2/S_269_ were detected in all the PBMC and tonsil samples (n = 12), while CD8^+^ T-cells directed at A2/Orf1ab_3183_ were found in only 33% of individuals (n = 12) and the lung tissues were uniformly negative (Fig. 3CiiD). Both the A2/S_269_^+^CD8^+^ and A2/Orf1ab_3183_^+^CD8^+^ were found over a broad range of ages (A2/S_269_: 5–68 years; A2/Orf1ab, 11–65 years). Moreover, the A2/S_269_^+^CD8^+^ T-cell frequency of 2.5×10^−6^ (mean, n = 12) in pre-COVID-19-healthy individuals was significantly lower than that found for COVID-19-exposed individuals (p = 0.0064; Fig. 3D). It thus seems that the A2/S_269_^+^CD8^+^ T-cells are indeed being activated and clonally expanded during SARS-CoV-2-infection. In contrast, there was no significant difference in frequencies for the A2/Orf1ab_3183_^+^CD8^+^ T-cells from the pre-pandemic and COVID-19 groups (p = 0.4121) (Fig 3D).

To further probe the the responsiveness of A2/SARS-CoV-2 CD8^+^ T-cells from uninfected versus convalescent COVID-19 donors, PBMCs or tonsil cells were stimulated with the S_269_ and Orf1ab_3183_ peptides and cultured *in vitro* for 10 days. In pre-pandemic ‘naïve’ subjects, no evidence of proliferation in culture was found for the A2/S_269_^+^CD8^+^ or A2/Orf1ab_3183_^+^CD8^+^ sets (Fig. 3E). In contrast, both the A2/S_269_^+^CD8^+^ and A2/Orf1ab_3183_^+^CD8^+^ T-cells from the COVID-19 donors increased significantly in numbers (*p* = 0.0357, Fig. 3E). Evidently the SARS-CoV-2/CD8^+^ T-cells from COVID-19 individuals (but not those from SARS-CoV-2 naïve subjects) were primed by SARS-CoV-2 and are thus, at least under *in vitro* conditions capable of clonal expansion.

### Activation profiles of SARS-CoV-2-specific A2/CD8^+^ T cells directly *ex vivo*

The activation profiles of A2/S_269_^+^CD8^+^ T-cells tested directly *ex vivo* from acute and convalescent patients were assessed by CD27, CD45RA and CD95 staining to determine the prevalence of the naïve (T_Naïve_) (CD27^+^CD45RA^+^CD95^−^), stem cell memory (T_SCM_) (CD27^+^CD45RA^+^CD95^+^), central memory (T_CM_)-like (CD27^+^CD45RA^−^), effector memory (T_EM_)-like (CD27^−^CD45RA^−^), and effector memory CD45RA (T_EMRA_) (CD27^−^CD45RA^+^) subsets (Fig 4A). Acute COVID-19 donors displayed the highest proportion (mean of 92%) EM of T_CM_-like A2/S_269_^+^CD8^+^ T-cells and a low proportion of T_EM_–like CD8^+^ T-cells. The A2/S_269_^+^ CD8^+^ T-cells from the convalescent versus acute subjects had a lower prevalence of T_CM_-like (mean of 50%) cells, and larger proportions of the T_Naïve_ (mean of 27%) and T_SCM_ (mean of 15%) sets, indicating that A2/S_269_^+^CD8^+^ T-cells expressing the optimally responsive T_CM_ phenotype fall off rapidly in blood sampled after the infection has resolved. Conversely, the majority of A2/S_269_^+^CD8^+^ T-cells within pre-pandemic children and adults were naïve(T_Naïve_; mean of 68% and 77%, respectively), while this subset was less prominent (mean of 46%) in the elderly. Interestingly older, uninfected people had a mean of 38% T_CM_-like A2/S_269_^+^CD8^+^ T-cells, similar to the frequency found for COVID-19 convalescents (mean of 50%), but less than that for IAV A2/M1_58_ (mean of 66%).

**Fig 4.**
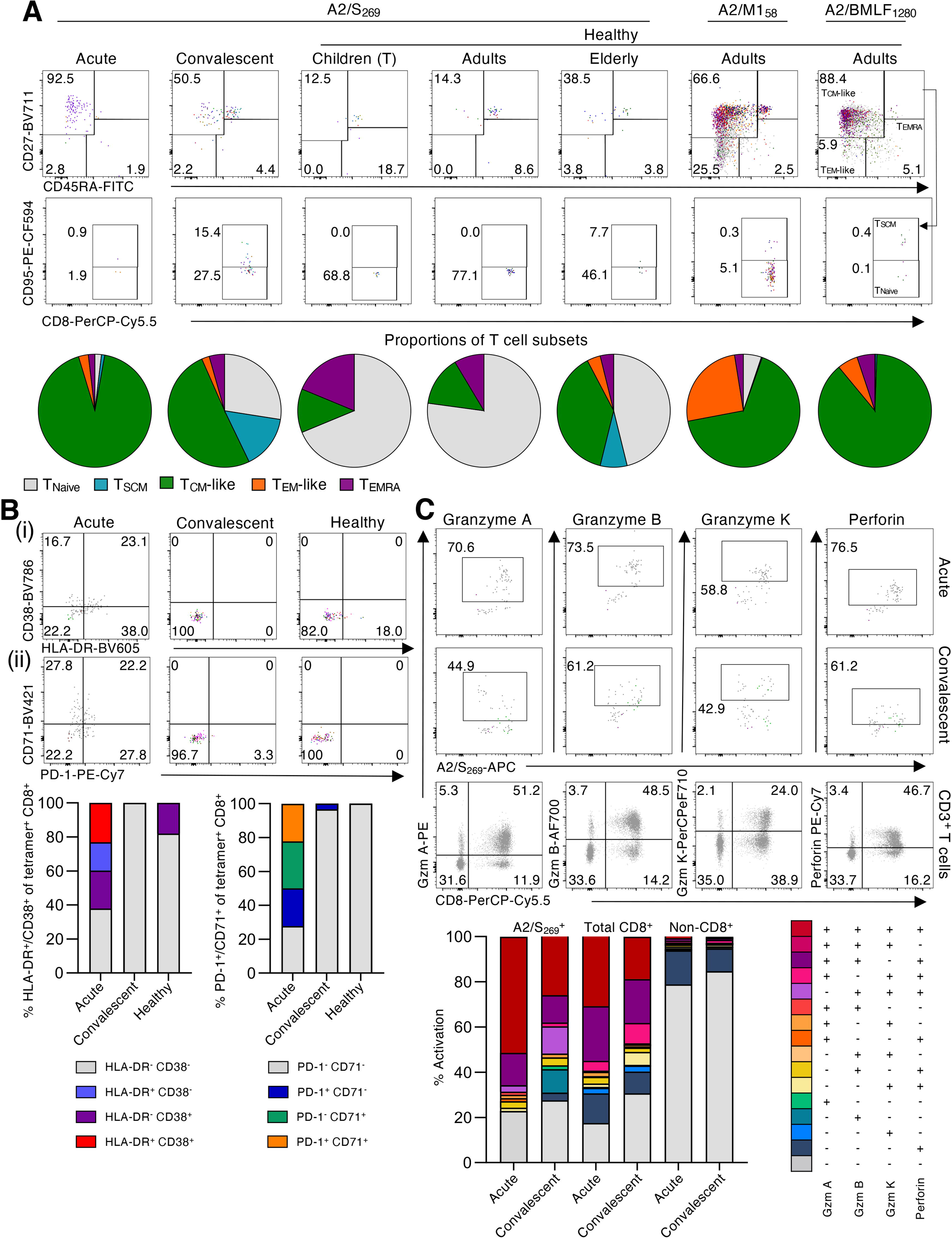
*Ex vivo* activation profiles of SARS-CoV-2-specific A2/CD8^+^ T-cells in COVID-19 subjects. (A) Overlayed FACS plots of A2/S_269_^+^CD8^+^ T-cells from acute COVID-19 *(n* = 3), convalescent COVID-19 *(n* = 11), healthy children (tonsils) *(n* = 4), healthy adults *(n* = 4) or healthy elderly donors *(n* = 4) show T_Naïve_ (CD27^+^CD45RA^+^CD95^−^), T_SCM_ (CD27^+^CD45RA^+^CD95^+^), T_CM_-like (CD27^+^CD45RA^−^). T_EM_-like (CD27^−^CD45RA^−^) and T_EMRA_ (CD27^−^CD45RA^+^) subsets. Pie charts display the proportion of each phenotype subset based on the combined data per each COVID-19 or healthy donor group. Overlayed FACS plots of A2/M1_58_^+^CD8^+^ and A2/BMLF_1280_^+^CD8^+^ T-cell memory phenotypes from healthy adults are also shown. (B) Overlayed FACS plots and combined frequencies of A2/S_269_^+^CD8^+^ T-cells showing (i) HLA-DR and CD38 or (ii) PD-1 and CD71 activation markers for acute *(n* = 3), convalescent *(n* = 11) and healthy donors *(n* = 12). (C) Overlayed FACS plots and combined frequencies of A2/S_269_^+^CD8^+^ T-cells showing granzyme A, B and K, and perforin staining for acute (*n* = 2) and convalescent (*n* = 3) donors. Representative FACS plots from one donor showing granzymes A, B, and K, and perforin of the total CD3^+^ T cell population. Combination gating was used to determine the frequency of cells with 1–4 functions for A2/S_269_^+^CD8^+^, total CD8^+^ or non-CD8^+^ T cells. Graphed data across multiple COVID-19 acute, COVID-19 convalescent or naïve subjects were combined for the activation and phenotypic analyses of A2/S_269_ CD8^+^ T-cells.

The expression profiles for HLA-DR, CD38, PD-1 and CD71 were also determined for tetramer^+^ A2/S_269_^+^CD8^+^ T-cells from the COVID-19 patients (Fig 4B). Only T cells from acutely-infected donors were positive for these activation markers, with the majority coming from one individual (COVID-19 #2). In contrast, the A2/S_269_^+^CD8^+^ T-cells from pre-pandemic and COVID-19 convalescent subjects were characterised by minimal levels of HLA-DR^+^CD38 and PD-1^+^CD71^−^ suggesting that, while the A2/S_269_^+^CD8^+^ set can be activated during the acute phase of the infection, it does not persist into short-term memory. Overall, our data suggest that naïve A2/SARS-CoV-2-specific CD8^+^ T-cells can indeed be expanded ∼5-fold and activated during the acute phase of COVID-19 but that, atypically for what we know for other readily resolved infections like influenza, both the extent of T-cell proliferation and the persistence of activated T-cells in the blood is low for (days 37–101 post disease onset) convalescent individuals.

To further investigate the suboptimal activation of SARS-CoV-2-specific CD8^+^ T-cells in COVID-19, the killing capacity of A2/S_269_^+^CD8^+^ T-cells was assessed by staining for granzyme A, B and K, and perforin directly *ex vivo*. Surprisingly, the majority of A2/S_269_^+^CD8^+^ T-cells at both acute (mean of 77.2%) and convalescent (mean of 72.4%) stages of COVID-19 expressed 3–4 cytotoxic granzymes/perforin (Fig 4C, Supplementary Fig 3), indicating their activation status. However, a similarly high expression level of granzymes/perforin was also found on the majority of total CD8^+^ T-cells (69–82.5%), as per our previous case report (13), but not on non-CD8^+^ T cells (mean of 15–21%). As it is highly unlikely that ∼80% of all CD8^+^ T cells in the peripheral blood during primary SARS-CoV-2 infection were antigen-specific (even if directed at several CD8^+^ T cell epitopes), this suggests that a high proportion of CD8^+^ T cells are activated via some ‘bystander’ mechanism during acute/convalescent COVID-19. The consequences, if any, of this effect for TCR-mediated activation merit further investigation.

## DISCUSSION

As the research community drives forward to design and evaluate novel vaccines and immunotherapies for COVID-19, concurrent efforts directed at understanding how immunity works in this disease process are largely focused on patient studies. Applying our established expertise in the analysis of T-cell-mediated immunity, we found here that the CD4^+^ ‘helper’ T-cell response looks relatively normal when compared with what happens in, for example, people who have been infected with an influenza A virus. However, when it comes to the virus-specific CD8^+^ T-cells that play an important role in ameliorating disease severity and driving recovery in other respiratory infections, our findings for COVID-19 are less encouraging. Though we were able to identify two SARS-CoV-2-specific CD8^+^ T-cell epitopes associated with the ubiquitous (in Caucasian) HLA-A*02:01 MHC-I glycoprotein (A2/S_269–277_ and A2/Orf1ab_3183–3191_) and found evidence for T-cell responsiveness, the results were not what we expected.

Our findings show that, while ‘early memory’ CD8^+^ T-cells can be detected in convalescent HLA-A*02:01 COVID-19 patients at frequencies ∼5-fold higher than those from pre-pandemic samples, the SARS-CoV-2-specific response was ∼10-fold lower than that found regularly for CD8^+^ T-cells directed at IAV or EBV epitopes. In general, there was an over-representation of SARS-CoV-2-specific tetramer^+^CD8^+^ T-cells expressing cell surface phenotypes that are considered to be characteristic of ‘stem cell memory’ and naïve precursor status, suggesting that the infectious process is in some way limiting both clonal expansion and differentiation of the ‘classical’ effector and central memory sets. An alternative explanation is, of course, that T-cell effectors are being generated but are localised to, and perhaps ‘consumed-in’ (driven to apoptosis?) sites of virus-induced pathology.

Even so, it is the case that SARS-CoV-2-specific CD8^+^ T-cells were found in all COVID-19 acute and convalescent donors, and in stored pre-pandemic PBMC and tonsil samples (but not lung tissues) from HLA-A*02:01 children, mature adults and the elderly. As the frequency of these naïve, pre-pandemic SARS-CoV-2-specific CD8^+^ T-cells (∼2.5×10^−6^) was numerically comparable to that found for naïve HIV (Gag_77–85_, SLYNTVATL), cancer (Survivin_96–104_) or HCV (NS3_1073_)-specific CD8^+^ T cell populations in healthy HLA-A*02:01^+^ individuals (19–21), both their presence and the fact that they were not readily expanded following *in vitro* stimulation suggests that they were not a product of prior exposure to some cross-reactive epitope. In fact, these are likely the naïve precursors that would be stimulated by appropriate prime-and-boost vaccine strategies.

Earlier experiments in a mouse model of SARS-CoV-1 showed that a conventional, CD8^+^ T-cell-targeted prime-and-boost approach indeed established substantial pools of memory SARS-CoV-1-specific CD8^+^ T-cells capable of driving protection against lethal SARS-CoV-1 infection (22). The fact that the frequencies of A2/S_269_^+^CD8^+^ T-cells in COVID-19 patients increased ∼5-fold suggest that these SARS-CoV-2-specific CD8^+^ T-cells proliferated to some extent during primary COVID-19, however not to the level of well-established memory CD8^+^ T-cell populations directed at other viral epitopes like IAV-specific A2/M1_58_ and EBV-specific A2/BMLF_1280_. Further studies are obviously needed to understand why this is so. In addition, as our acquaintance with this novel CoV continues, we will be able to determine if there is long-term survival (at least at > 1 year) of SARS-CoV-2-specific CD8^+^ memory T-cells following primary COVID-19 along with whether, in now healthy survivors, they can be activated and clonally expanded following challenge with an appropriate vaccine.

Surprisingly, the memory A2/S_269_^+^CD8_269_^+^ T-cell populations in convalescent subjects were dominated by stem cell memory, central memory and naïve phenotypes, and lacked expression of the CD38, HLA-DR, PD-1 and CD71 activation markers. This is in stark contrast to the highly activated T_EM_ and T_EMRA_ profiles found *ex vivo* in both short-term (day 25) and long-term (7 months) memory A2/M1_58_^+^CD8^+^ T-cells following avian A/H7N9 influenza infection (23, 24). These minimal activation profiles for epitope-specific CD8^+^ T-cells in early COVID-19 convalescence could possibly reflect suboptimal priming of A2/S_269_^+^CD8^+^ T-cells in primary COVID-19. Furthermore, a recent study by Zhou *et al* (2020) demonstrated perturbed dendritic cell and T cell function in SARS-CoV2 infection (25). Impaired dendritic cell function might negatively impact antigen processing and presentation to CD8^+^ T cells, thus at least partially explaining the limited differentiation of SARS-CoV-2-specific CD8^+^ T cells observed here.

It remains unclear whether this is broadly representative of primary CD8^+^ T-cell responses in COVID-19 or specific to the epitopes analysed here. There is a possibility that there are other HLA-A*02:01-restricted immunodominant epitopes, or even immunodominant epitopes restricted by other HLAs in HLA-A*02:01^+^ COVID-19 patients. The A2/S_269_ epitope identified in our study was, however, also independently reported in a recent pre-print (26), suggesting it is a common HLA-A*02:01 epitope. Moreover, it is also possible that CD8^+^ T cells directed towards other HLA-A*02:01-restricted epitopes might have expressed high levels of PD-1 and thus had an impaired capacity to expand *in vitro* due to their exhausted phenotype. Further identification of CD8^+^ T-cell epitopes across a broad range of HLA class I alleles and SARS-CoV-2 proteins is needed to provide a more detailed landscape of CD8^+^ T-cell responses in COVID-19, their *ex vivo* frequencies and activation profiles. In-depth analysis of epitope-specific T-cell responses in severe and critical cases is also essential if we are to understand whether the activation profiles of early CD8^+^ T-cell memory reflect disease severity. And, as the range of candidate vaccines that are tested through phase 1 trials expands, it would also be of great benefit to determine whether the characteristics of memory CD8^+^ T-cells generated in the absence of active infection look more optimal than those described here.

Stimulation with overlapping peptides led to the expansion of SARS-CoV-2-specific CD8^+^ and CD4^+^ T-cells *in vitro*, although CD4^+^ T-cells dominated the response. This might support, at least partially, the previous elegant study showing that CD4^+^ T-cells but not CD8^+^ T-cells were of a greater importance in primary SARS-CoV-1 infection, as depletion of CD4^+^ T-cells (but not CD8^+^ T-cells) led to delayed viral clearance from the lungs, associated with reduced neutralizing antibody and cytokine production (27). It is also important to note that the Spike peptide pool from Miltenyi Biotec used here spans only selected regions (304–338, 421–475, 492–519, 683–707, 741–770, 785–802 and 885–1273) rather than the entire protein, thus some CD8^+^ and CD4^+^ T-cell responses could have been missed. Recent evidence revealed that Th2 and Th17 cytokine profiles in COVID-19 patients can be associated with differential disease outcomes (28). Our analyses focused on Th1 cytokine responses for CD4^+^ T cells, leaving Th2 and Th17 cytokine responses unknown. Different cytokine profiles of epitope-specific CD4^+^ T-cells should be investigated in future studies, especially when SARS-CoV2-specific CD4^+^ T cell epitopes are identified.

Our early report on immunity to COVID-19 in one of Australia’s first patients suggested that broad and concomitant immune responses were associated with recovery from mild-to-moderate COVID-19 disease (13). The key immune populations detected included antibody-secreting cells, helper follicular T-cells, activated (CD38^+^HLA-DR^+^) CD8^+^ and CD4^+^ T-cells, together with progressive increases in SARS-CoV-2-specific IgM and IgG antibodies. Subsequent studies confirmed the activation of both CD4^+^ and CD8^+^ T-cells as indicated by cell-surface marker expression (14, 15). The present, much more extensive yet focused analysis does, however, raise questions concerning the integrity of the epitope-specific CD8^+^ T cell response in COVID-19. Given the variation in disease outcome with this infection, that obviously merits much more detailed analysis.

## Data Availability

All the data are available upon request

## ACKNOWLEDGMENTS

We thank all the participants involved in the study, Robyn Esterbauer, Hannah Kelly, Jane Batten and Helen Kent for support with the cohort. We thank Jill Garlick, Janine Roney, Anne Paterson and the research nurses at the Alfred Hospital. This work was supported by the Clifford Craig Foundation to KLF and KK, NHMRC Leadership Investigator Grant to KK (1173871), NHMRC Program Grant to DLD (#1132975), Research Grants Council of the Hong Kong Special Administrative Region, China (#T11–712/19-N) to KK, the Victorian Government (SJK, AKW), MRFF award (#2002073) to SJK and AKW, MRFF Award (#1202445) to KK, NHMRC program grant 1149990 (SJK), NHMRC project grant 1162760 (AKW). AWC is supported by a NHMRC Career Development Fellowship (#1140509), KK by NHMRC Senior Research Fellowship (1102792), DLD by a NHMRC Principal Research Fellowship (#1137285) and SJK by NHMRC Senior Principal Research Fellowship (#1136322). JR is supported by an ARC Laureate fellowship. JRH is supported by the Melbourne Research Scholarship from The University of Melbourne. CES has received funding from the European Union’s Horizon 2020 research and innovation program under the Marie Skłodowska-Curie grant agreement (#792532). LH is supported by the Melbourne International Research Scholarship (MIRS) and the Melbourne International Fee Remission Scholarship (MIFRS) from The University of Melbourne. JAJ is supported by an NHMRC Early Career Fellowship (ECF) (#1123673).

## METHODS

### Study participants and ethics statement

35 subjects were recruited into this study. Acute and convalescent COVID-19 were recruited via the Alfred Hospital, University of Melbourne or James Cook University. Seven of the donors were admitted to hospital during their active infection (Supplementary Table 1). Acute COVID-19 cases were admitted to the hospital ward, with two patients requiring oxygen support (Supplementary Table 1). Healthy donors were recruited via University of Melbourne or buffy packs obtained from the Australian Red Cross LifeBlood (West Melbourne, Australia) (Supplementary Table 2). Tonsils were obtained from healthy individuals undergoing tonsillectomy (Tasmania, Australia). Lung samples were obtained prior to the COVID-19 pandemic via the Alfred Hospital’s Lung Tissue Biobank. All blood and tonsil donors were HLA typed by VTIS (Melbourne, Australia). Peripheral blood was collected in heparinized tubes and peripheral blood monocular cells (PBMCs) were isolated via Ficoll-Paque separation.

Experiments conformed to the Declaration of Helsinki Principles and the Australian National Health and Medical Research Council Code of Practice. Written informed consents were obtained from all blood donors prior to the study. Lung tissues were obtained from deceased organ donors after written informed consents from the next of kin. Written informed consents were obtained from participants’ parents or guardians for underage tonsil tissue donors. The study was approved by the Alfred Hospital (#280/14), The University of Melbourne (#2056689, #2056761, #1442952, #1955465, and #1443389), the Australian Red Cross Lifeblood (ID 2015#8), the Tasmanian Health and Medical (ID H0017479) and the James Cook University (H7886) Human Research Ethics Committees.

### Cell lines and reagents

C1R.A*02:01 cells were maintained in RF-10 medium (RPMI-1640 with 10% heat-inactivated FCS (Gibco; Thermo Fisher Scientific)), with 0.3 mg/mL hygromycin-B (Life Technologies). Overlapping synthetic peptides spanning the entire Nucleocapsid (N) and Membrane (M) proteins, and selected regions of SARS-CoV2 Spike (S) were purchased from Miltenyi Biotec (PepTivator SARS-CoV-2 Prot_N (130–126–699), Prot_M (130–126–703) and Prot_S (130–126–701)) and reconstituted in 80% DMSO. Synthetic SARS-CoV2 peptides predicted to bind HLA-A*02:01 were purchased from GenScript and reconstituted in DMSO. Tetramers were generated from soluble, biotinylated HLA-A2 monomers. Briefly, HLA α-heavy chain with C-terminal BirA biotinylation motif and β2-microglobulin were expressed and purified as inclusion bodies in *E. coli*, solubilized in 6 M Guanidine HCl and refolded with either S_269_ or Orf1ab_3183_ peptide, in buffer containing 50 mM Tris pH8, 3 M urea, 0.4 M Arginine, 2 mM oxidised Glutathione, 20 mM Glutathione, 2 mM EDTA, 10 mM PMSF and cOmplete™ protease inhibitor (Roche). Following dialysis in 10 mM Tris, HLA monomers were purified via DEAE and HiTrapQ ion exchange chromatography, and biotinylated with BirA ligase in 50 mM Bicine pH 8.3, 10 mM ATP, 10 mM magnesium acetate and 100 µm d-biotin. Following S200 gel permeation chromatography fully biotinylated HLA monomers were stored at –80 °C and conjugated to fluorescently-labeled streptavidins (SA), PE-SA or APC-SA (BD Biosciences) at an 8:1 monomer to SA molar ratio to form pMHC-I tetramers.

### Intracellular cytokine staining (ICS)

PBMCs and tonsil samples were stimulated with either 0.6 nmol of overlapping SARS-CoV2 peptides or 1 µM A2/SARS-CoV2 predicted peptides for 10 days in RF-10 medium (+20 U/mL IL-2) (29). On d10, cells were stimulated with peptides for 6 hrs in the presence of GolgiPlug and GolgiStop (BD Bioscience) plus 10 U/mL IL-2, and the SARS-CoV2-reactive T cells quantified using anti-IFN-_γ_-V450, anti-TNF-AF700, anti-MIP-1β-APC, anti-CD107a-AF488 (BD) and anti-perforin-PE-Cy7 (BioLegend) ICS (30). CD8^+^ T cells specific for A2/SARS-CoV2 epitopes were quantified using IFN-_γ_/TNF ICS with C1R.A*02:01 cells used as antigen-presenting cells.

### *Ex vivo* tetramer enrichment and phenotypic analysis

Cells (1–86×10^6^) were stained with A2/S_269_-PE and A2/Orf1ab_3183_-APC tetramers at 1:100 for 1 hr in MACS buffer (PBS with 0.5% BSA and 2 mM EDTA). PBMCs and tonsil samples were incubated with anti-PE and anti-APC microbeads (Miltenyi) and tetramer^+^ cells were enriched using magnetic separation (18, 19). Cells were stained with anti-CD71-BV421, anti-CD4-BV650, anti-CD27-BV711, anti-CD38-BV786, anti-CD14-APC-H7, anti-CD19-APC-H7, anti-CD45RA-FITC, anti-CD8-PerCP-Cy5.5, anti-CD95-PE-CF594 and anti-PD1-PE-Cy7 (BD) and anti-CD3-BV510, anti-HLA-DR-BV605 (Biolegend) and Live/Dead near-infrared (Invitrogen) for 30 mins, washed, resuspended in MACS buffer and analysed by flow cytometry. Lung cells were stained with the following alterations; anti-CD69-BV421, anti-CD103-FITC (BioLegend), and anti-CD45RO-PE-Cy7 (Thermo Fisher Scientific). To detect granzymes and perforin, cells were stained with A2/S_269_-APC and enriched, then surface stained with anti-CD27-BV711, anti-CD3-PE-CF594, anti-CD4-BV650, anti-CD45RA-FITC, anti-CD19-APC-H7, anti-CD14-APC-H7 (BD), anti-CD8-BV421

(Biolegend) and Live/Dead aqua (Invitrogen) and stained intracellularly with anti-Granzyme-B-AF700 (BD), anti-Perforin-PE-Cy7 (Biolegend), anti-Granzyme-A-PE and anti-Granzyme-K-PerCP-efluor710 (eBiosceince) after fixation/permeabilization with eBioscience^TM^ Foxp3/Transcription Factor Staining Buffer Set (Thermo Fisher Scientific). Samples were acquired on a BD LSRII Fortessa. Flow cytometry data were analyzed using FlowJo v10 software.

---

**Supp Fig 1**. (A) Representative FACS plots of IFN-_γ_/TNF staining of CD4^+^ and CD8^+^ T-cells stimulated with PMA-I (positive control) and DMSO (negative control) after d10 expansion *in vitro* with SARS-CoV-2 overlapping peptide pools. (B) Representative FACS plots after d10 culture with A2/SARS-CoV-2 predicted peptides. IFN-_γ_/TNF staining of CD8^+^ T-cells stimulated with PMA-I (positive control), DMSO (negative control), SARS-CoV-2 predicted peptide pool and individual A2/SARS-CoV2 peptides.

**Supp Fig 2**. (A) Representative FACS plots of A2/S_269_^+^CD8^+^ and A2/Orf1ab_3183_^+^CD8^+^ T-cells from unenriched, enriched and flow-through samples of (i) convalescent and (ii) acute COVID-19 PBMCs. (B) Representative FACS plots of A2/M1_58_^+^CD8^+^ and A2/BMLF_1280_^+^CD8^+^ T-cells from unenriched, enriched and flow-through samples of healthy PBMCs. (C) Representative FACS plots of A2/S_269_^+^CD8^+^ and A2/Orf1ab_3183_^+^CD8^+^ T-cells from unenriched, enriched and flow-through samples of adult and elderly PBMCs, and children tonsils (T).

## REFERENCES

1. E. Dong, H. Du, L. Gardner, An interactive web-based dashboard to track COVID-19 in real time. Lancet Infect Dis 20, 533–534 (2020).

2. Z. Wu, J. M. McGoogan, Characteristics of and Important Lessons From the Coronavirus Disease 2019 (COVID-19) Outbreak in China: Summary of a Report of 72 314 Cases From the Chinese Center for Disease Control and Prevention. Jama 10.1001/jama.2020.2648 (2020).

3. Y. Hu et al., Prevalence and severity of corona virus disease 2019 (COVID-19): A systematic review and meta-analysis. J Clin Virol 127, 104371 (2020).

4. E. K. Stokes et al. (2020) Coronavirus Disease 2019 Case Surveillance — United States, January 22–May 30, 2020. (MMWR Morb Mortal Wkly Rep 2020), pp 759–765.

5. H. Meng et al., CT imaging and clinical course of asymptomatic cases with COVID-19 pneumonia at admission in Wuhan, China. J Infect 81, e33–e39 (2020).

6. Q.-X. Long et al., Antibody responses to SARS-CoV-2 in patients with COVID-19. Nat Med 26, 845–848 (2020).

7. J. Zhao et al., Antibody responses to SARS-CoV-2 in patients of novel coronavirus disease 2019. Clin Infect Dis 10.1093/cid/ciaa344 (2020).

8. Y. Wang et al., Kinetics of viral load and antibody response in relation to COVID-19 severity. J Clin Invest 10.1172/JCI138759 (2020).

9. F. Wu et al., Neutralizing antibody responses to SARS-CoV-2 in a COVID-19 recovered patient cohort and their implications. medRxiv 10.1101/2020.03.30.20047365, 2020.2003.2030.20047365 (2020).

10. N. Le Bert et al., SARS-CoV-2-specific T cell immunity in cases of COVID-19 and SARS, and uninfected controls. Nature 10.1038/s41586-020-2550-z (2020).

11. J. Zhao et al., Recovery from the Middle East respiratory syndrome is associated with antibody and T cell responses. Sci Immunol 2, eaan5393 (2017).

12. J. Mateus et al., Selective and cross-reactive SARS-CoV-2 T cell epitopes in unexposed humans. Science 10.1126/science.abd3871, eabd3871 (2020).

13. I. Thevarajan et al., Breadth of concomitant immune responses prior to patient recovery: a case report of non-severe COVID-19. Nat Med 26, 453–455 (2020).

14. A. Grifoni et al., Targets of T Cell Responses to SARS-CoV-2 Coronavirus in Humans with COVID-19 Disease and Unexposed Individuals. Cell 181, 1489–1501.e1415 (2020).

15. D. Weiskopf et al., Phenotype and kinetics of SARS-CoV-2-specific T cells in COVID-19 patients with acute respiratory distress syndrome. Sci Immunol 5, eabd2071 (2020).

16. H. Y. Zheng et al., Elevated exhaustion levels and reduced functional diversity of T cells in peripheral blood may predict severe progression in COVID-19 patients. Cell Mol Immunol 17, 541–543 (2020).

17. F. Amanat et al., A serological assay to detect SARS-CoV-2 seroconversion in humans. Nat Med 10.1038/s41591-020-0913-5 (2020).

18. S. A. Valkenburg et al., Molecular basis for universal HLA-A*0201–restricted CD8+ T-cell immunity against influenza viruses. Proc Natl Acad Sci 10.1073/pnas.1603106113, 201603106 (2016).

19. T. H. Nguyen et al., Understanding CD8(+) T-cell responses toward the native and alternate HLA-A*02:01-restricted WT1 epitope. Clin Transl Immunology 6, e134 (2017).

20. E. J. Grant et al., Lack of Heterologous Cross-reactivity toward HLA-A*02:01 Restricted Viral Epitopes Is Underpinned by Distinct T Cell Receptor Signatures. J Biol Chem 291, 24335–24351 (2016).

21. K. Kedzierska, M. Koutsakos, The ABC of Major Histocompatibility Complexes and T Cell Receptors in Health and Disease. Viral Immunol 33, 160–178 (2020).

22. R. Channappanavar, C. Fett, J. Zhao, D. K. Meyerholz, S. Perlman, Virus-specific memory CD8 T cells provide substantial protection from lethal severe acute respiratory syndrome coronavirus infection. J Virol 88, 11034–11044 (2014).

23. Z. Wang et al., Recovery from severe H7N9 disease is associated with diverse response mechanisms dominated by CD8+ T cells. Nat Commun 6, 6833 (2015).

24. Z. Wang et al., Clonally diverse CD38+HLA-DR+CD8+ T cells persist during fatal H7N9 disease. Nat Commun 9, 824 (2018).

25. R. Zhou et al., Acute SARS-CoV-2 Infection Impairs Dendritic Cell and T Cell Responses. Immunity 10.1016/j.immuni.2020.07.026 (2020).

26. A. S. Shomuradova et al., SARS-CoV-2 epitopes are recognized by a public and diverse repertoire of human T-cell receptors. medRxiv 10.1101/2020.05.20.20107813, 2020.2005.2020.20107813 (2020).

27. J. Chen et al., Cellular immune responses to severe acute respiratory syndrome coronavirus (SARS-CoV) infection in senescent BALB/c mice: CD4+ T cells are important in control of SARS-CoV infection. J Virol 84, 1289–1301 (2010).

28. C. Lucas et al., Longitudinal analyses reveal immunological misfiring in severe COVID-19. Nature 10.1038/s41586-020-2588-y (2020).

29. M. Koutsakos et al., Human CD8+ T cell cross-reactivity across influenza A, B and C viruses. Nat Immunol 20, 613–625 (2019).

30. E. B. Clemens et al., Towards identification of immune and genetic correlates of severe influenza disease in Indigenous Australians. Immunol Cell Biol 94, 367–377 (2016).

